# SARS-CoV-2 antigen and antibody prevalence among UK staff working with cancer patients during the COVID-19 pandemic

**DOI:** 10.1101/2020.09.18.20197590

**Authors:** David M Favara, Karen McAdam, Anthony Cooke, Alex Bordessa-Kelly, Ieva Budriunaite, Sophie Bossingham, Sally Houghton, Rainer Doffinger, Nicola Ainsworth, Pippa G Corrie

## Abstract

**Background:** International guidelines for testing potentially immunosuppressed cancer patients receiving non-surgical anticancer therapies for SARS-CoV-2 (COVID-19) are currently lacking. The value of routinely testing staff treating cancer patients is not known.

**Methods:** Patient-facing oncology department staff at work during the COVID-19 pandemic consented to have a nasopharyngeal swab SARS-CoV-2 antigen test by polymerase chain reaction (PCR) and blood tests for SARS-CoV-2 antibody using a laboratory Luminex-based assay and a rapid point-of-care (POC) assay on 2 occasions 28 days apart in June and July 2020.

**Findings:** 434 participants were recruited: nurses (58·3%), doctors (21·2%), radiographers (10·4%) and administrators (10·1%). 82% were female; median age 40-years (range 19-66). 26·3% reported prior symptoms suggestive of SARS-CoV-2 infection and 1·4% tested PCR-positive prior to June 2020. All were PCR-negative at both study day 1 and 28. 18·4% were SARS-CoV-2 sero-positive on day 1 by Luminex, of whom 42·5% also tested positive by POC. 47·5% of Luminex sero-positives had antibodies to both nucleocapsid (N) and surface (S) antigens. Nurses (21·3%) and doctors (17·4%) had higher prevalence trends of Luminex sero-positivity compared with administrators (13·6%) and radiographers (8·9%) (*p*=0.2). 38% of sero-positive participants reported previous symptoms suggestive of SARS-CoV-2 infection, a 1·9-fold higher odds than sero-negative participants (*p*=0·01). 400 participants re-tested on day 28: 13·3% were Luminex sero-positive of whom 92·5% were previously positive and 7·5% newly positive. Nurses (16·5%) had the highest seroprevalence trend amongst staff groups (*p*=0·07). 32·5% of day 1 sero-positives became sero-negative by day 28: the majority being previously reactive to the N-antigen only (*p*<0·0001).

**Interpretation:** The high prevalence of SARS-CoV-2 IgG sero-positivity in oncology nurses, and the high decline of positivity over 4 weeks supports regular antigen and antibody testing in this staff group for SARS-CoV-2 as part of routine patient care prior to availability of a vaccine.

**Funding:** ACT, NHS

**Evidence before this study:** To identify studies involving oncology healthcare workers and SARS-CoV-2 exposure during the COVID-19 pandemic, we searched PubMed and Medrxiv for articles published between January 1 and July 31 using the following search terms “COVID-19”, “SARS-CoV-2”, “oncology staff”, “healthcare workers” without language restriction. To date, no large study has specifically reported and tracked patient-facing oncology staff SARS-CoV-2 exposure.

**Added value of this study:** To the best of our knowledge, this is the first study specifically investigating SARS-CoV-2 exposure in UK patient-facing oncology staff who were at work during the peak of the COVID-19 pandemic between March and June 2020. 18·4% of staff were SARS-CoV-2 antibody positive at the start of June 2020 suggesting prior SARS-CoV-2 infection, while 32·5% of those antibody-positive cases became antibody-negative 28 days after the first sample collection. The highest seroprevalence rates at both time points were recorded in nurses.

**Implications of all the available evidence:** These results justify incorporating SARS-CoV-2 PCR and antibody testing of oncology nurses into international guidelines for managing cancer patients treated with non-surgical anticancer treatments prior to availability of a functional vaccine.

## Introduction

The global SARS-CoV-2 (COVID-19) pandemic has caused substantial morbidity and economic turmoil across the world. Despite ongoing research endeavours, an effective treatment or vaccine remains elusive. Various populations in multiple countries have been tested for SARS-CoV-2 infection by polymerase chain reaction (PCR) to detect viral antigen, with a wide range of reported asymptomatic positive carrier rates (4-80%).^1-5^ Antibody tests are the gold standard for providing evidence of exposure to pathogens following an adaptive immune response, including SARS-CoV-2 infection.^6,7^ Although reports of immunoglobulin (Ig)M and IgG antibody production rates against SARS-CoV-2 in COVID-19 pandemic survivors has varied, the first large Chinese series (n=285) of hospitalised SARS-CoV-2 PCR-positive patients reported that 100% of them developed SARS-CoV-2 IgG within 19 days of symptom onset.^8^ Another study showed that 511 (82%) of 624 SARS-CoV-2 PCR-positive out-patients developed antibodies 10-14 days after symptom onset.^9^ In a small French study of 14 SARS-CoV-2 PCR-positive healthcare workers (HCWs), 10 (71%) developed detectable SARS-CoV-2 antibodies 15 days after PCR testing.^10^

The prevalence of SARS-CoV-2 sero-positivity among asymptomatic HCWs not working directly with SARS-CoV-2 infected hospital in-patients has not been extensively investigated. Current data suggests that the sero-positivity rate may be very low, as evidenced by a French study in which only 3 (1·3%) of 230 asymptomatic PCR-naïve HCWs were SARS-CoV-2 IgG-positive. All 3 HCWs were PCR-negative at the time of testing, suggesting past exposure with successful eradication of the virus.^10^ A Spanish study reported a similarly low SARS-CoV-2 sero-positive rate in asymptomatic hospital staff (11/578; 1·9%).^11^ Data from screening asymptomatic HCWs for active SARS-CoV-2 viral infection is also limited, but again suggest very low rates. In the UK, one study showed a peak asymptomatic HCW infection rate of 7·1% in late March 2020, decreasing to 1·1% five weeks later^12^, whilst another study reported that 30 (3%) of 1,032 asymptomatic hospital HCWs tested in April 2020 were SARS-CoV-2 PCR-positive.^13^

People with cancer may be more likely to contract SARS-CoV-2 infection.^14,15^ This risk is amplified by multiple hospital attendances required for diagnosis, treatment and follow-up. Although recent results suggest that anti-cancer treatment does not increase mortality in infected cancer patients^16^, it may increase the risk of serious complications following infection^14,15^, so guidance is needed to safeguard both patients and oncology staff. There is currently no data regarding oncologist-specific SARS-CoV-2 infection and immunity rates within the UK, while risk of viral transmission between HCWs working with cancer patients is not known. The ‘COVID-19 Serology in Oncology Staff’ (CSOS) study was a multi-centre UK study aiming to provide an indication of staff infection rates in order to inform national guidance on managing cancer patients treated within secondary care non-surgical oncology departments, which is currently lacking. The CSOS study investigated SARS-CoV-2 serology (IgG) and asymptomatic viral carriage in patient-facing oncology staff who worked clinically during the peak of the UK COVID-19 pandemic, with sample collection at multiple time points.^17^ The primary objective was to determine the percentage of SARS-CoV-2 sero-positive oncology staff in the workplace during the pandemic. Secondary outcomes included the proportion of previously symptomatic and asymptomatic SARS-CoV-2 sero-positivity, how long SARS-CoV-2 sero-positivity lasted, the rate of persistent asymptomatic PCR-positivity over time, and the proportion of people who did not become sero-positive following a positive PCR result.

## Methods

We recruited staff involved in treating cancer patients in non-surgical oncology departments at three UK secondary care hospitals in the Eastern Region: the Queen Elizabeth Hospital Kings Lynn NHS Foundation Trust (QEH), a 515-bed hospital serving 331,000 people; North West Anglia NHS Foundation Trust (Peterborough City Hospital, NWA), a 635-bed hospital serving 700,000 people; and Cambridge University Hospitals NHS Foundation Trust (CUH), a 1400-bed academic hospital and cancer centre serving 1,300,000 people. Staff were excluded if they had not been working during the pandemic peak from March–June 2020, or were not patient-facing. Participants had to be working within the oncology department ward or out-patient setting and not primarily within a dedicated SARS-CoV-2 in-patient ward. Staff who returned to work after self-isolating due to SARS-CoV-2 symptoms, or exposure to a potentially affected household member (as per UK government rules) were eligible to participate. Following consent, samples were collected during June 2020 (day 1 samples) and July 2020 (day 28 samples). Samples collected were a nasopharyngeal swab for SARS-CoV-2 PCR testing and blood for SARS-COV-2 antibody testing. Anonymised participant characteristics were collected: age, sex, job role, smoking status, history of any underlying health condition, details of any suspected SARS-CoV-2 illness/exposure and leave taken, dates of start to resolution of presumed or confirmed SARS-CoV-2 illness, date of any prior SARS-CoV-2 tests and results. Regulatory approval for the study was granted by the UK Health Research Authority (IRAS: 284231; 26/5/2020). Data was analysed using Prism 8 (Graphpad Software) and R. One-way ANOVA was used to compare multiple means; Fisher’s exact test to compare groups and categorical variables; and the Mann-Whitney U test to compare day 1 antibody-positive outcomes at day 28.

### SARS-CoV-2 PCR assay

RNA extraction: Zymo (Cambridge, UK) Quick-RNA 96 kit (R1053). SARS-CoV-2 PCR detection performed using Primerdesign’s (Eastleigh, UK) Coronavirus COVID-19 Genesig RT-PCR assay (Z-Path-COVID-19-CE) and a Roche LightCycler 480 in a 96-well format. All kits used according to manufacturer guidelines.

### SARS-CoV-2 serology assays

Two different antibody assays were used to detect SARS-CoV-2 antibodies: a rapid point-of-care (POC) finger-prick assay and a laboratory-based assay. POC test: Abbexa (Cambridge, UK) COVID-19 IgG/IgM Rapid Test Kit (abx294171) detecting antibodies against the SARS-CoV-2 nucleocapsid (N) and spike (S)-antigens. Manufacturer claimed no cross-reactivity with antibodies against other coronaviruses (HKU1, OC43, NL63, 229E) and a sensitivity of 98·5% and specificity 97·94%. Results were read 10-15 minutes after assaying.

The laboratory-based assay was a SARS-CoV-2 IgG multiplex particle-based flow cytometry (Luminex) assay developed at CUH detecting antibodies against the SARS-CoV-2 N and full-length S-antigens^18^, requiring 4mL of blood in a serum tube. Sensitivity and specificity were determined at 84% and 100% respectively, using a cohort of pre-pandemic healthy controls versus a cohort of unselected SARS-CoV2 PCR-positive patients. At the time of writing, testing for cross-reactivity against other coronaviruses had not been performed for this semi-quantitative assay. A level of detection comparison was performed between the above-mentioned SARS-CoV-2 IgG assays using sequentially-diluted SARS-CoV-2 IgG-positive and negative sera.

## Results

434 oncology department staff were recruited (Supplementary Figure 1). Participant characteristics are summarised in Table 1 (aggregated) and Supplementary Table 1.1 (by hospital site).The majority of participants were nurses (253/434; 58·3%) and doctors (92/434; 21·2%), with smaller proportions of radiographers (45/434; 10·4%) and administrative staff (44/434; 10·1%). Physiotherapists (n=8) were grouped with the nurses since their job role involved similar close physical patient contact. No radiographers were recruited at QEH as this site had no radiotherapy facility (Supplementary Table 1). The majority of participants were female (356/434; 82%) (*p*<0.001) and median participant age was 41 (range 19-66) years. Administrators had the highest median age (median 47.5 years) followed by doctors (42 years), nurses (39 years) and radiographers (33 years) (*p*<0.001). Chronic underlying health conditions were reported in 103/434 (23·7%) of all participants (Table 1), with asthma and hypertension being the most common in all groups (Supplementary table 1.2).

**Table 1.** Participant characteristics.

|  | Total | Doctors | Nurses | Radiographers | Administrators | *P* value |
| --- | --- | --- | --- | --- | --- | --- |
| **Number** | 434 | 92 | 253 | 45 | 44 |  |
| **Mean age (median; range)** | 41 (40; 19-66) | 43.1 (42; 25-63) | 40.4 (39; 21-65) | 35.1 (33; 19-57) | 45.5 (47.5; 24-66) | <0.0001 |
| **Female** | 356/434 (82%) | 49/92 (53.3%) | 227/253 (89.7%) | 39/45 (86.7%) | 41/44 (93.2%) | <0.0001 |
| **Male** | 78/434 (18%) | 43/92 (46.7%) | 26/253 (10.3%) | 6/45 (13.3%) | 3/44 (6.8%) |  |
| **Smoker** | 37/434 (8.5%) | 2/92 (2.1%) | 22/253 (8.7%) | 2/45 (4.4%) | 11/44 (25%) | 0.0003 |
| **Underlying health condition** | 103/434 (23.7%) | 15/92 (16.3%) | 60/253 (23.7%) | 14/45 (31.1%) | 14/44 (31.8%) | 0.1 |

114 (26·3%) participants reported symptoms suggestive of SARS-CoV-2 infection prior to study entry (Table 2); 33·7% of doctors, 25·7% of nurses, 22·7% of administrators and 17·8% of radiographers reported symptoms. Mean duration of reported symptoms was 11·8 (median 7·5; range 1-61) days and was similar for all staff groups (*p*=0·1). The mean time from symptom resolution to the first study sample collection date was 68·5 days (median 67; range 1-172), with no differences between staff groups (*p*=0·8). 178 (41%) participants underwent nasopharyngeal PCR testing for SARS-CoV-2 infection between March and June 2020 prior to initiation of this study (doctors having the highest frequency (*p*=0·006)): 70 were symptomatic (70/114; 61·4% of the symptomatic population) and 108 were asymptomatic (tested as part of hospital screening programmes). 6/70 (12·9%) of the tested symptomatic population were SARS-CoV-2 PCR-positive (3 doctors and 3 nurses), whilst the asymptomatic group were all PCR-negative. The majority of participants (320; 73·7%) reported no prior symptoms. Of these, 47 (14·7%) reported a household member having symptoms suggestive of SARS-CoV-2 infection.

**Table 2.** Results. (NA – not applicable)

|  | Total | Doctors | Nurses | Radiographers | Administrators | *p* value |
| --- | --- | --- | --- | --- | --- | --- |
| **Previously symptomatic:** |  |  |  |  |  |  |
| Prior symptoms suggestive of COVID-19 | 114/434 (26.3%) | 31/92 (33.7%) | 65/253 (25.7%) | 8/45 (17.8%) | 10/44 (22.7%) | 0.2 |
| Mean symptom duration in days (median; range) | 11.8 (7.5; 1-61) | 14.5 (7; 1-61) | 10.9 (8; 1-48) | 9.5 (7; 2-20) | 11.3 (8; 1-37) | 0.1 |
| Mean time (days) from symptom resolution to sample collection (median; range) | 68.5 (67; 1-172) | 68.3 (70; 1-169) | 70.2 (67; 8-172) | 67.1 (63; 28-139) | 59.4 (69; 12-94) | 0.8 |
| Pre-study SARS-CoV-2 PCR test | 178/434 (41%) | 46/92 (50%) | 108/253 (42.7%) | 15/45 (33.3%) | 9/44 (20.5%) | 0.006 |
| Pre-study positive PCR test | 6/178 (3.4%) | 3/46 (6.5%) | 3/108 (2.7%) | 0/15 | 0/9 | 0.6 |
| **Previously asymptomatic:** |  |  |  |  |  |  |
| Asymptomatic during pandemic | 320/434 (73.7%) | 61/92 (66.3%) | 188/253 (74.3%) | 37/45 (82.2%) | 34/44 (77.3%) | 0.2 |
| Asymptomatic, but household member possibly symptomatic | 47/320 (14.7%) | 11/61 (18%) | 27/188 (14.4%) | 3/37 (8.1%) | 6/34 (17.6%) | 0.7 |
| **DAY 1: Study sample collection:** |  |  |  |  |  |  |
| Positive SARS-CoV-2 PCR test | 0/434 | 0/92 | 0/253 | 0/45 | 0/44 |  |
| Positive SARS-CoV-2 IgG (Luminex test) | 80/434 (18.4%) | 16/92 (17.4%) | 54/253 (21.3%) | 4/45 (8.9%) | 6/44 (13.6%) | 0.2 |
| *Luminex test positive for antibodies to both N and S antigens* | 38/80 (47.5%) | 8/16 (50%) | 28/54 (51.9%) | 0/4 | 2/6 (33.3%) | 0.2 |
| *Luminex test positive for antibodies to only N antigen* | 32/80 (40%) | 7/16 (43.8.5%) | 18/54 (33.3%) | 3/4 (75%) | 4/6 (66.7%) | 0.2 |
| *Luminex test positive for antibodies to only S antigen* | 10/80 (12.5%) | 1/16 (6.3%) | 8/54 (14.8%) | 1/4 (25%) | 0/6 | 0.5 |
| Positive SARS-CoV-2 IgG (POC test) | 34/434 (7.8%) | 8/92 (8.7%) | 22/253 (8.7%) | 0/45 | 4/44 (9.1%) | 0.2 |
| **Luminex test Subgroup analyses: Day 1** |  |  |  |  |  |  |
| Sero-positive after pre-study positive PCR | 6/6 (100%) | 3/3 (100%) | 3/3 (100%) | NA | NA |  |
| Sero-positive, previously symptomatic | 30/114 (26.3%) | 9/31 (29%) | 19/65 (29.2%) | 0/8 (0%) | 2/10 (20%) | 0.4 |
| Sero-positive, previously asymptomatic | 50/320 (15.6%) | 7/61 (11.5%) | 35/188 (18.6%) | 4/37 (10.8%) | 4/34 (11.7%) | 0.4 |
| Sero-positive, previously asymptomatic with possible household contact | 12/47 (25.5%) | 2/11 (18.2%) | 10/27 (37%) | 0/3 (0%) | 0/6 (0%) | 0.2 |
| Sero-positive, with chronic underlying health condition | 18/103 (17.5%) | 4/15 (26.7%) | 12/60 (20%) | 2/14 (14.3%) | 0/14 (0%) | 0.2 |
| Sero-positive, smokers | 9/37 (24.3%) | 1/2 (50%) | 6/22 (27.3%) | 0/2 (0%) | 2/11 (18.2%) | 0.6 |
| **DAY 28: Study sample collection:** | **n=400** | **n=82** | **n=237** | **n=38** | **n=43** |  |
| Positive SARS-CoV-2 nasopharyngeal PCR swab | 0/400 | 0/82 | 0/237 | 0/38 | 0/43 |  |
| Positive SARS-CoV-2 IgG (Luminex test) | 53/400 (13.3%) | 9/82 (10.9%) | 39/237 (16.5%) | 1/38 (2.6%) | 4/43 (9.3%) | 0.07 |
| *Luminex test positive for antibodies to both N and S antigens* | 34/53 (64.2%) | 7/9 (77.8%) | 25/39 (64.1%) | 0/1 (0%) | 2/4 (50%) | 0.5 |
| *Luminex test positive for antibodies to only N antigen* | 9/53 (16.9%) | 0/9 (0%) | 7/39 (17.9%) | 0/1 (0%) | 2/4 (50%) | 0.2 |
| *Luminex test positive for antibodies to only S antigen* | 10/53 (18.9%) | 2/9 (22.2%) | 7/39 (17.9%) | 1/1 (100%) | 0/4 (0%) | 0.3 |
| Sero-positive on days 1 and 28 (Luminex test) | 49/53 (92.5%) | 9/9 (100%) | 35/39 (89.7%) | 1/1 (100%) | 4/4 (100%) | 0.4 |
| Sero-negative on day 1, sero-positive on day 28 (Luminex test) | 4/53 (7.5%) | 0/9 (0%) | 4/39 (10.3%) | 0/1 (0%) | 0/4 (0%) | 0.4 |
| Sero-positive on day 1, sero-negative on day 28 (Luminex test) | 26/80 (32.5%) | 5/16 (29.4%) | 17/54 (31.2%) | 2/4 (50%) | 2/6 (33.3%) | 0.4 |
| Total Positive SARS-CoV-2 IgG POC test) | 29/400 (7.3%) | 5/82 (6.1%) | 22/237 (9.3%) | 0/38 (0%) | 2/43 (4.7%) | 0.2 |
| Sero-positive on days 1 and 28 (POC test) | 25/29 (86.2%) | 5/5 (100%) | 18/22 (81.8%) | NA | 2/2 (100%) | 0.7 |
| Sero-negative on day 1, sero-positive on day 28 (POC test) | 4/29 (13.8%) | 0/5 (0%) | 4/22 (18.2) | NA | 0/2 (0%) | 0.7 |
| Sero-positive on day 1, sero-negative on day 28 (POC test) | 6/34 (17.6%) | 1/8 (12.5%) | 3/22 (13.6%) | NA | 2/4 (50%) | 0.2 |

All participants tested at day 1 (June 2020) were nasopharyngeal swab PCR-negative for SARS-CoV-2 (Table 2 and Supplementary Table 2). SARS-CoV-2 IgG was detected at day 1 in 80 (18·4%) participants using the Luminex test, and in 34 (7·8%) participants using the POC test (Table 2). All sero-positive participants using the POC test were also sero-positive using the Luminex test. The Luminex test was able to detect SARS-CoV-2 IgG at 1:100 dilution, which the POC could not (Supplementary Table 3), so results from the Luminex assay were used to define participant serology status (sero-positive or sero-negative).

At day 1, nurses were the staff group with the highest proportion of SARS-CoV-2 sero-positivity (54/253; 21·3%), followed by doctors (16/92; 17·4%), administrators (6/44; 13·6%) and radiographers (4/45; 8·9%), although these differences were not statistically significant (*p*=0·2) (Table 2). Of the 80 sero-positive participants, 38 (47·5%) had antibodies to both the nucleocapsid (N) and surface (S) SARS-CoV-2 antigens, 32 (40%) had antibodies only to the N-antigen and 10 (12·5%) had antibodies only to the S-antigen (Table 2). All participants with a previous positive nasopharyngeal PCR result tested sero-positive (6/6; 100%) with detectable antibodies to both N and S SARS-CoV-2 antigens.

37·5% (30/80) of day 1 sero-positive participants reported previous symptoms consistent with SARS-CoV-2 infection during the pandemic: a 1·9-fold higher odds than sero-negative participants (84/354; 23·7%) (*p*=0·01) (Table 2). Out of the total number of participants reporting previous symptoms, 50/320 (15·6%) had detectable SARS-CoV-2 antibodies at day 1, which was similar across staff groups *(p=*0·4). Out of 47 participants who had no prior symptoms but had been exposed to a suspected infected household member, 12 (25·5%) had positive antibodies. These findings are visualised in Figure 1. There was no significant difference in sero-positivity between those with (18/103; 17·5%) or without (62/331; 18·7%) underlying health conditions (*p*=0·2), or between smokers (9/37; 24·3%) and non-smokers (71/397; 17·9%) (*p*=0.6) (Table 2).

**Figure 1.**
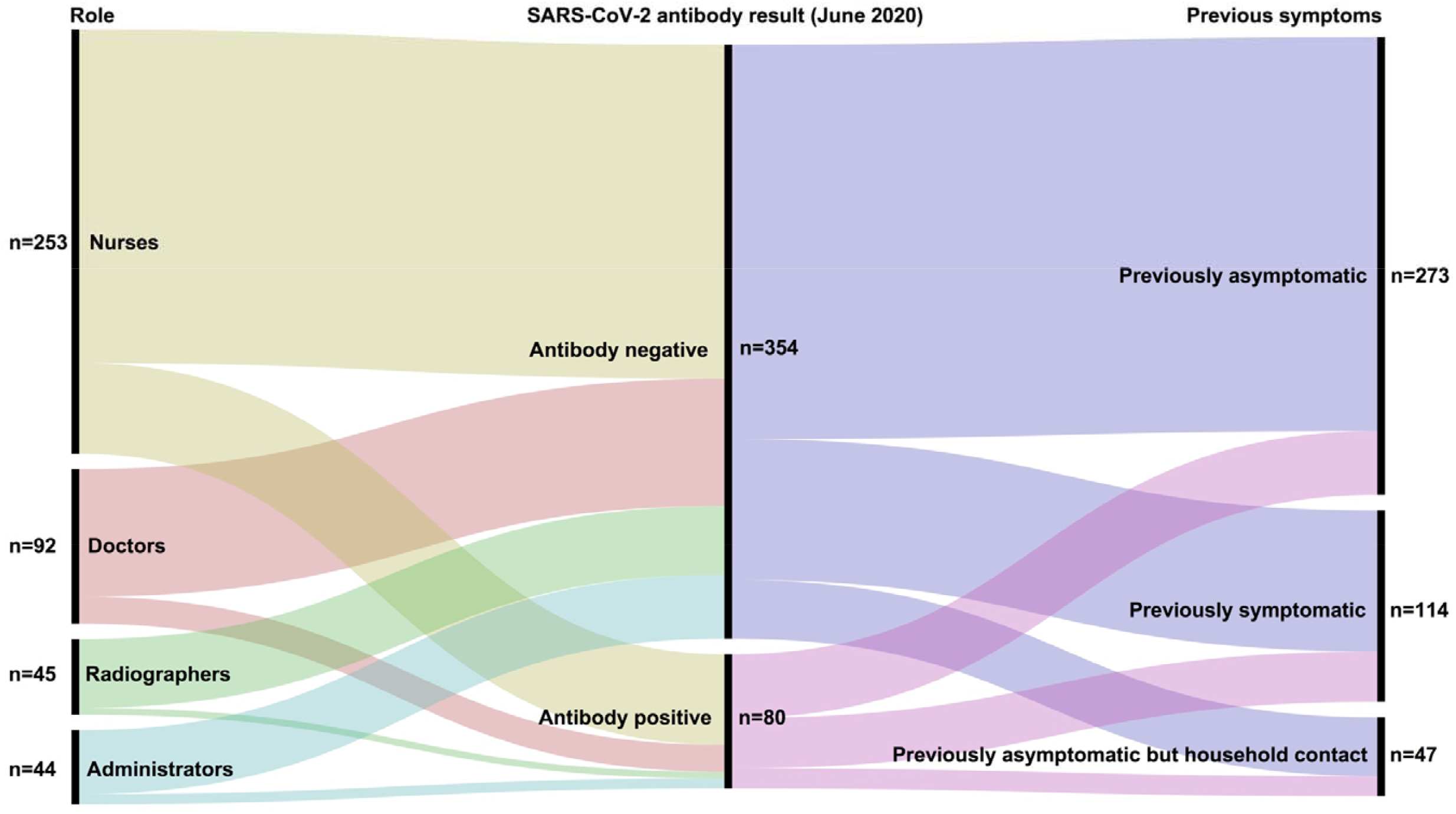
Summary of relationship between role, previous symptoms and antibody results (June 2020 sample collection). All participants were nasopharyngeal swab SARSCOV-2 PCR negative at time of SARS-COV-2 antibody testing.

400 (92·2%) participants were retested on day 28 during July 2020 with 34 (7·8%) lost to follow-up (Table 2). All participants remained nasopharyngeal swab PCR-negative for SARS-CoV-2 on retesting. Positive SARS-CoV-2 serology was detected in 53/400 (13·3%) participants using the Luminex test, and in 29/400 (7·3%) using the POC test. Of the 29 sero-positives detected by the POC test at day 28, 27 were also positive with the Luminex test, but 2 were not. The 2 discordant results were previously negative using both assays at day 1 (Table 2 and Supplementary Table 2) with both participants denying any intervening symptoms or virus exposure between testing dates.

Nurses remained the staff group with the highest rate of Luminex test sero-positivity (39/237; 16·5%), followed by doctors (9/82; 10·9%), administrators (4/43; 9·3%) and radiographers (1/38; 2·6%), although the trend did not reach statistical significance (*p*=0·07).

32.5% (26/80) of those previously sero-positive at day 1 with the Luminex test were sero-negative on day 28, whilst 5/80 (6·25%) were lost to follow-up (Table 2). Of those participants who were sero-positive on day 28, 49/53 (92·5%) were persistently positive on both day 1 and day 28, and 4/53 (7·5%) were new-seroconversions with no reported symptoms. All those with a positive SARS-CoV-2 PCR test prior to study entry remained SARS-CoV-2 sero-positive using the Luminex test (6/6; 100%).

Of the 53 sero-positive participants identified at day 28 with the Luminex test, 34 (64·2%) had antibodies to both the N and S SARS-CoV-2 antigens, whilst 9 (16·9%) had antibodies to only the N-antigen, and 10/53 (18·9%) to only the S-antigen (Table 2). Loss of sero-positivity affected all antigen-target groups, with the largest decrease affecting those who were N-antigen positive on day 1 (*p*<0·0001) (Table 2 and Supplementary Table 4). At day 28, 3 of the 33 participants originally positive on day 1 with antibodies reactive to the N-antigen were lost to follow-up, whilst 18 became sero-negative and 2 lost their N-antigen antibodies and developed S-antigen antibodies, signifying a 68·9% reduction in positivity rate (Supplementary Table 4). This loss only affected those who were previously asymptomatic (Supplementary Table 4). These findings are visualised in Figure 2. None of the previously reported SACRS-CoV-2 PCR-positive participants became sero-negative by day 28. Luminex and POC results from both day 1 and 28 are shown in Table 2 (aggregated) and Supplementary Table 2 (by hospital site).

**Figure 2.**
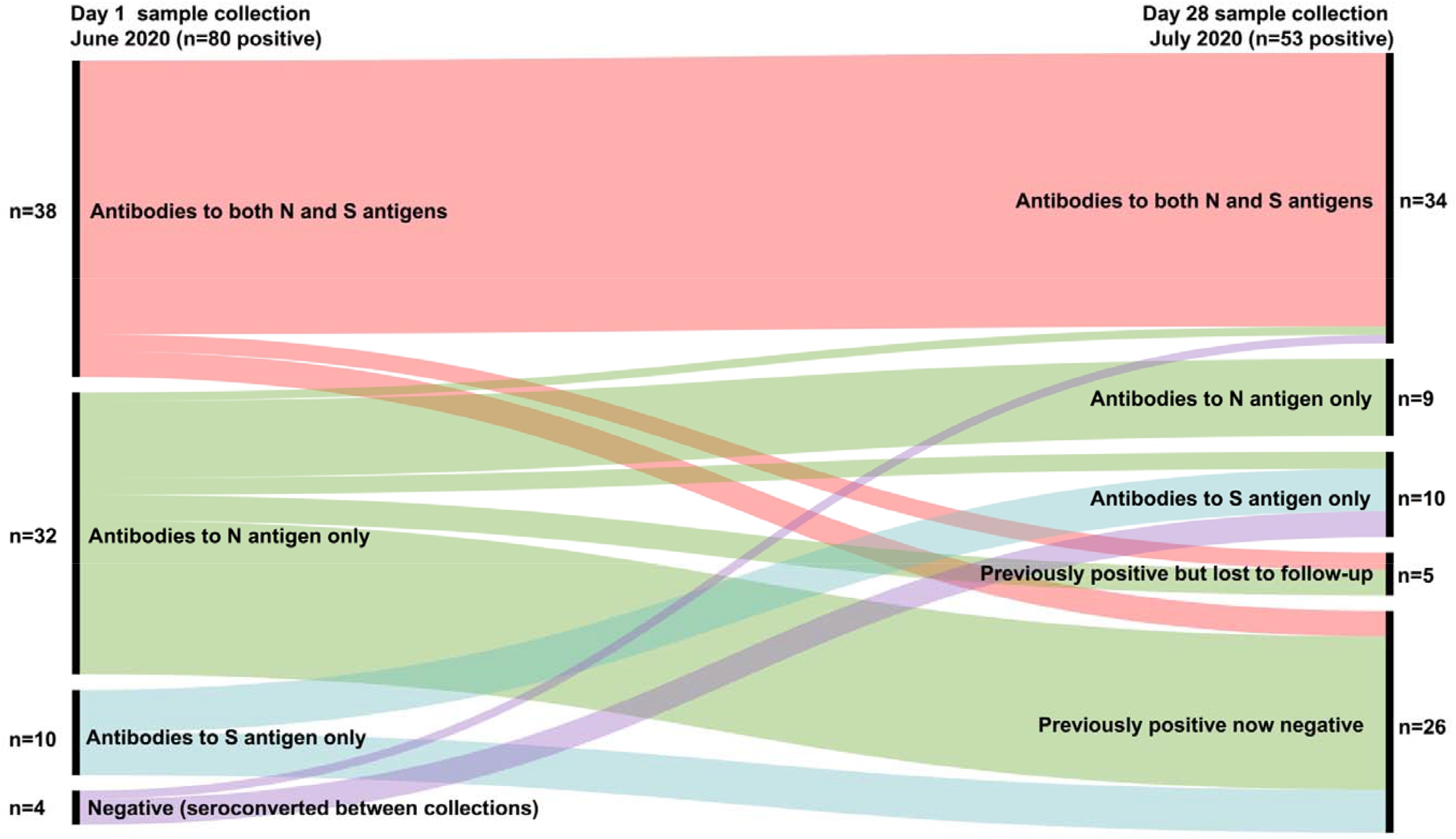
Summary of Relationship between day 1 and day 28 positive antibody results (by SARS-CoV-2 antigen target).

## Discussion

The CSOS study is the first report of SARS-CoV-2 exposure in patient-facing staff working within non-surgical oncology departments in 3 hospitals of differing size and structure located in the Eastern region of the UK. Staff working during the COVID-19 pandemic peak between March and June 2020 were tested for both SARS-CoV-2 antigen and antibodies. All participants were SARS-CoV-2 PCR-negative at both testing time-points 28 days apart.

We used 2 different antibody detection methods to compare and contrast their potential clinical utility for measuring SARS-CoV-2 serology. The POC test, with the advantage of immediate read-out within 15 minutes, was reported by the manufacturer to have high sensitivity and specificity and to not cross-react with the 4 main other coronavirus types. The Luminex test, which had a processing time of 2-3 hours, but real-time results returned between 2-4 days, was able to detect antibodies at a much lower concentration level compared with the POC test. Although already being used for routine clinical practice at one of the study hospitals (CUH), at the time of writing, the Luminex test remains in development and has not been investigated for cross-reactivity against other coronaviruses, so we cannot exclude the possibility of false-positives and that participants may have been exposed to other coronaviruses rather than SARS-CoV-2 infection. In a recent report, 35% of 44 healthy donors demonstrated memory T-cell reactivity to S-antigen exposure, suggesting cross-reactivity from past encounters with endemic coronaviruses^19^. In contrast, a study investigating a coronavirus antigen microarray containing antigens from SARS-CoV-2, SARS-CoV, MERS-CoV and endemic human coronaviruses (such as the common cold) found that there was no cross-reactivity between antibodies against SARS-CoV-2’s N-antigen and antigens of endemic coronaviruses or MERS-CoV (N-antigen cross-reactivity was found only between SARS-CoV-2 and SARS-CoV)^20^. If we had only used the POC test, we would have missed over half of cases with lower levels of sero-positivity.

Using the Luminex test, 1 in 5 staff members were SARS-CoV-2 sero-positive despite being PCR-negative at the time of sampling, which suggests a substantial past exposure and infection rate, especially considering that the majority of participants were not working within dedicated SARS-COV-2 wards, or participating in high-risk aerosol-generating procedures in known SARS-CoV-2 patients. Similarly, our finding of a 15·6% asymptomatic infection rate (evidenced by sero-positivity in June 2020) is higher than large-scale community seroprevalence studies which have reported sero-positive rates ranging from 5% (in 51,958 Spanish participants)^21^ to 6% (in 109,076 UK participants)^22^ and in studies of HCW groups ranging from 5·4% (in 244 French HCWs)^10^ to 9.3% (in 578 Spanish HCWs)^11^. Our finding that symptomatic individuals had a higher odds of being sero-positive than asymptomatic individuals fits with previous reports^23^. We hypothesise that most staff became infected at work rather than in the community, due to the nature of the hospital admission process and general patient care, compounded by the issue of personal protective equipment (PPE) being less readily available within the NHS in the early stages of the pandemic.

This hypothesis is further substantiated when considering outcomes amongst staff groups. Nurses appear to be the most affected group followed by doctors, administrators and radiographers: while nurses represented 58% of total participants, they also accounted for 67% of total sero-positive cases on day 1 and 74% of total sero-positive cases on day 28. The seroprevalence rate of 21·3% of nurses tested on day 1 and 16·5% tested on day 28 was the highest of all the 4 staff groups. Although not statistically significant compared to other staff groups (due to the smaller numbers of other staff groups recruited), the daily interactions of nurses with multiple patients at close quarters will undoubtedly contribute to these stark statistics.

Our finding that 32·5% of sero-positive participants became seronegative during our 4-week study interval is consistent with emerging reports showing declining sero-positivity over time. These include a Chinese study reporting 26·2% of 61 participants becoming seronegative 8 weeks after documented SARS-CoV-2 infection,^24^ a UK study reporting declining neutralising antibodies in 65 participants sequentially sampled up to 94 days after onset of SARS-CoV-2 symptoms,^25^ and an American study reporting that, in a cohort of 37 participants, the SARS-CoV-2 antibody half-life was 37 days.^26^ Our finding that asymptomatic participants were the majority of those who became sero-negative during the 4-week testing interval fits the previously mentioned Chinese study which reported that 40% of 30 asymptomatic participants became sero-negative 8 weeks after initial diagnosis, compared with 12·9% of 31 symptomatic participants, suggesting that SARS-CoV-2 antibody response is dependent on disease severity.^24^ This has important implications for HCWs, as it remains unclear whether low-level or decreasing SARS-CoV-2 antibodies provide adequate protection from re-infection. In this regard, recent data regarding the neutralizing ability of SARS-CoV-2 antibodies is promising, as well as the potential protective role of memory cytotoxic T-cells in individuals with past mild or asymptomatic infection, which suggest that SARS-CoV-2 immunity may rely on multiple mechanisms rather than only humoral immunity.^9,27^ Recently, an Icelandic study reported that 1107/1215 (91%) of its previously SARS-CoV-2 PCR-positive participants were found to be sero-positive, remaining so for 120 days.^28^ It remains possible that our and other published studies reporting declining sero-positivity over time were confounded by assay-related detection weaknesses, however all were performed using high sensitivity and specificity laboratory methods in current clinical use. Another possibility is that sustained SARS-CoV-2 antibody production is dependent on a number of factors including the dose of virus received, host genetics including immune system genotype and phenotype, and the strain of virus present in that geographic location.

Interestingly, the mean time of 68.5 days from symptom resolution to study antibody testing suggests that we may have missed many participants who could have been asymptomatic but infected and whose SARS-CoV-2 antibody levels had decreased to sero-negative levels before we commenced sample collection. Our finding over the 28-day study period that 2 previously asymptomatic sero-positive participants lost their previously detectable IgG antibodies against the N-antigen whilst developing detectable IgG antibodies against the S-antigen is novel. This suggests an evolving humoral immune response in both participants, which was also noted in other day 1 sero-positive participants who manifested increased levels of anti-SARS-CoV-2 antibodies on day 28. IgG antibodies targeting SARS-COV-2 S-antigen have been reported to elicit both neutralising^29,30^ and non-neutralising responses.^31^ Additionally, N-antigen exposure has been reported to elicit a strong and viable immune response.^31^ A previous report found that detection of N-antigen was more sensitive than S-antigen in detecting early SARS-CoV-2 infection.^32^

The strengths of this study are its multi-site structure, concurrent use of 2 different antibody assays and concurrent PCR-testing over 2 different time-points. Participant drop-out rate between the 2 time-points was low, suggesting high motivation to remain involved. The study will continue collecting samples at intervals until a vaccine becomes wide-available. This will enable assessment of the durability of SARS-CoV-2 antibody responses in previously symptomatic and asymptomatic individuals, as well as the impact of any future surges in infection rates that may occur in the coming months.

This study sets the first sero-positivity baseline for UK oncology staff and provides new information to consider incorporating into international guidance on safeguarding patients due to start or having started non-surgical anticancer treatments. Currently, the most comprehensive guidance comes from the UK^33^ and recommends that all patients receiving systemic anticancer therapy in the UK should undergo SARS-CoV-2 PCR-testing prior to starting treatment and considerations should be given to subsequent testing at intervals during treatment. The guidance advises testing HCWs in the broadest sense. We propose that there should be a focus on routinely testing oncology nursing staff for both SARS-CoV-2 antigen and antibodies until an effective vaccine comes available. Testing antibody status alongside viral carriage status on a large scale would enable the SARS-CoV-2 antibody dynamics to be better understood. Ours and other study findings^24,26^ that sero-positivity fluctuates within 4 weeks suggests that testing should be performed at least monthly (ideally weekly PCR-testing with fortnightly serology), although this recommendation remains open to debate. Increasing availability of lower-cost, high sensitivity and specificity SARS-CoV-2 testing methods should make this targeted approach viable, would help protect patients and staff and enable containment and tracking of new, asymptomatic infections, should they occur. The recent report of an asymptomatic individual re-infected with a phylogenetically distinct SARS-CoV-2 strain 4.5 months after first asymptomatic infection suggests that SARS-CoV-2 may continue spreading despite populations acquiring herd immunity.^34^ In this specific case, viral sequencing and homology comparisons showed that the second virus had amino acid changes in its S-antigen, allowing for immune evasion leading to re-infection.^34^ It remains unknown whether this individual had a detectable antibody response following their first infection, but routine longitudinal serology testing could help answer these intriguing questions.

## Data Availability

Data available following reasonable request.

## Acknowledgements

The authors would like to thank Mandy Coventry, Paula Santos, Pran Parsheni (Oncology Department, Queen Elizabeth Hospital Kings Lynn), Helen Bowyer, Holly Warman, Terri-Anne Baker, Kerrie Cavanah (Oncology Research Department, Peterborough City Hospital), Doreen Milne, Catherine Booth (Cambridge Clinical Trials Unit, Addenbrooke’s Hospital), Tracy Cook, Justine Kane, Dr Lynsey Drewett (Cambridge Breast Cancer Research Unit, Addenbrooke’s Hospital), Cherry Publica, Cara Caasi (Cambridge Clinical Trials Unit, Addenbrooke’s Hospital), Dr Sara Lightowler and Agata Zielinska for help with sample collection; Dr Hilary Martin (Wellcome Sanger Institute and University of Cambridge) and Dr Mireia Crispin Ortuzar (CRUK Cambridge Institute, University of Cambridge) for their critical readings of the manuscript; Mark Cross for fundraising for the Oncology Research Fund at Peterborough City Hospital; and Oorvanshi Joshi and Jakub Letowski (Laboratory Research Services, Addenbrooke’s Hospital).

## Author contributions

DMF designed the study and wrote the manuscript. PGC, NA and KA contributed to critical revision of the study and manuscript and read and approved the final version. RD, SH, IB and SB processed the Luminex SARS-CoV-2 assays, and AC processed the SARS-CoV-2 PCR assays. AB assisted with sample collection. All authors read and approved the manuscript.

---

**Supplementary figure 1.**
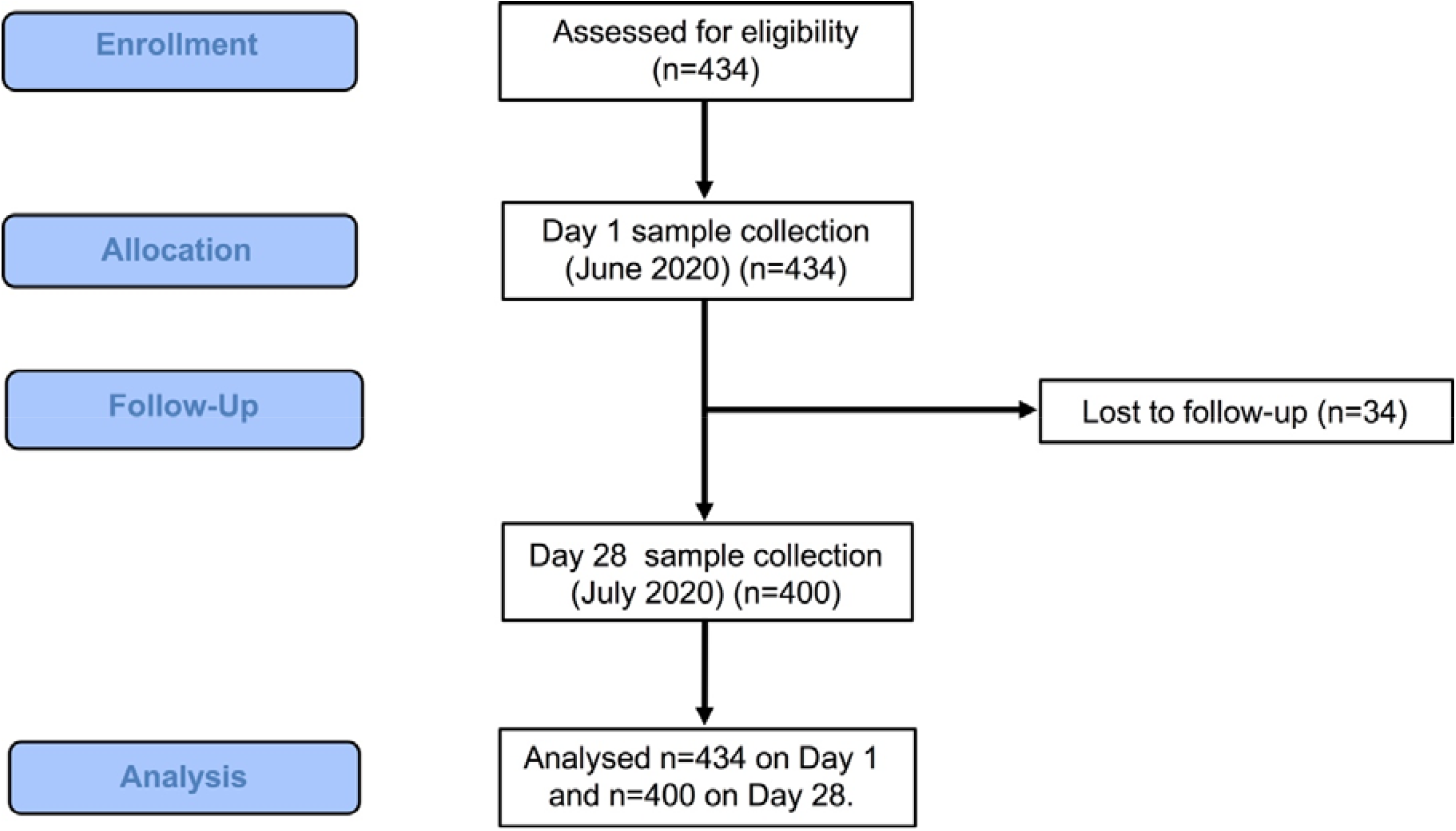
Study outline.

**Supplementary Table 1.1.** Participant characteristics by participating hospital site. (NA – not applicable)

| Site 1: QEH | Total | Doctors | Nurses | Radiographers | Administrators |
| --- | --- | --- | --- | --- | --- |
| **Number** | 70 | 15 | 45 | 0 | 10 |
| **Mean age (median; range)** | 42 (41; 23-64) | 41 (42; 25-60) | 40 (40; 23-61) | NA | 48 (49; 25-64) |
| **Female** | 61/70 (87.1%) | 9/15 (60%) | 42/45 (93.3%) | NA | 10/10 (100%) |
| **Male** | 9/70 (12.9%) | 6/15 (40%) | 3/45 (6.7%) | NA | 0/10 (%) |
| **Smoker** | 7/70 (10%) | 0/15 (0%) | 5/45 (11.1%) | NA | 2/10 (20%) |
| **Underlying health condition** | 16/70 (22.9%) | 2/15 (13.3%) | 9/45 (20%) | NA | 5/10 (50%) |

| Site 2: NWA | Total | Doctors | Nurses | Radiographers | Administrators |
| --- | --- | --- | --- | --- | --- |
| **Number** | 97 | 10 | 68 | 6 | 13 |
| **Mean age (median; range)** | 42.74 (42; 25-62) | 45.6 (44; 32-57) | 41.2 (39; 26-62) | 47 (48.5; 37-57) | 46.9 (48; 25-36) |
| **Female** | 86/97 (88.7%) | 6/10 (60%) | 63/68 (92.6%) | 6/6 (100%) | 11/13 (8.5%) |
| **Male** | 11/97 (11.3%) | 4/10 (40%) | 5/68 (7.4%) | 0 | 2/13 (15.4%) |
| **Smoker** | 8/97 (8.2%) | 0 | 3/68 (4.4%) | 0 | 5/13 (38.5%) |
| **Underlying health condition** | 33/97 (34%) | 3/10 (30%) | 22/68 (32.3%) | 3/6 (50%) | 5/13 (38.5%) |

| Site 3: CUH | Total | Doctors | Nurses | Radiographers | Administrators |
| --- | --- | --- | --- | --- | --- |
| **Number** | 267 | 67 | 140 | 39 | 21 |
| **Mean age (median; range)** | 40.1 (39; 19-66) | 43.2 (42; 30-63) | 40.1 (39.5; 21-65) | 33.3 (31; 19-38) | 43.5 (42; 24-42) |
| **Female** | 209/267 (78.3%) | 34/67 (50.7%) | 122/140 (87.1%) | 33/39 (84.6%) | 20/21 (95.2%) |
| **Male** | 58/267 (22.7%) | 33/67 (49.3%) | 18/140 (12.8%) | 6/39 (15.4%) | 1/21 (4.8%) |
| **Smoker** | 22/267 (8.2%) | 2/67 (2.9%) | 14/140 (10%) | 2/39 (5.1%) | 4/21 (19%) |
| **Underlying health condition** | 54/267 (20.2%) | 10/67 (14.9%) | 29/140 (20.7%) | 11/39 (28.2%) | 4/21 (19%) |

**Supplementary Table 1.2.**
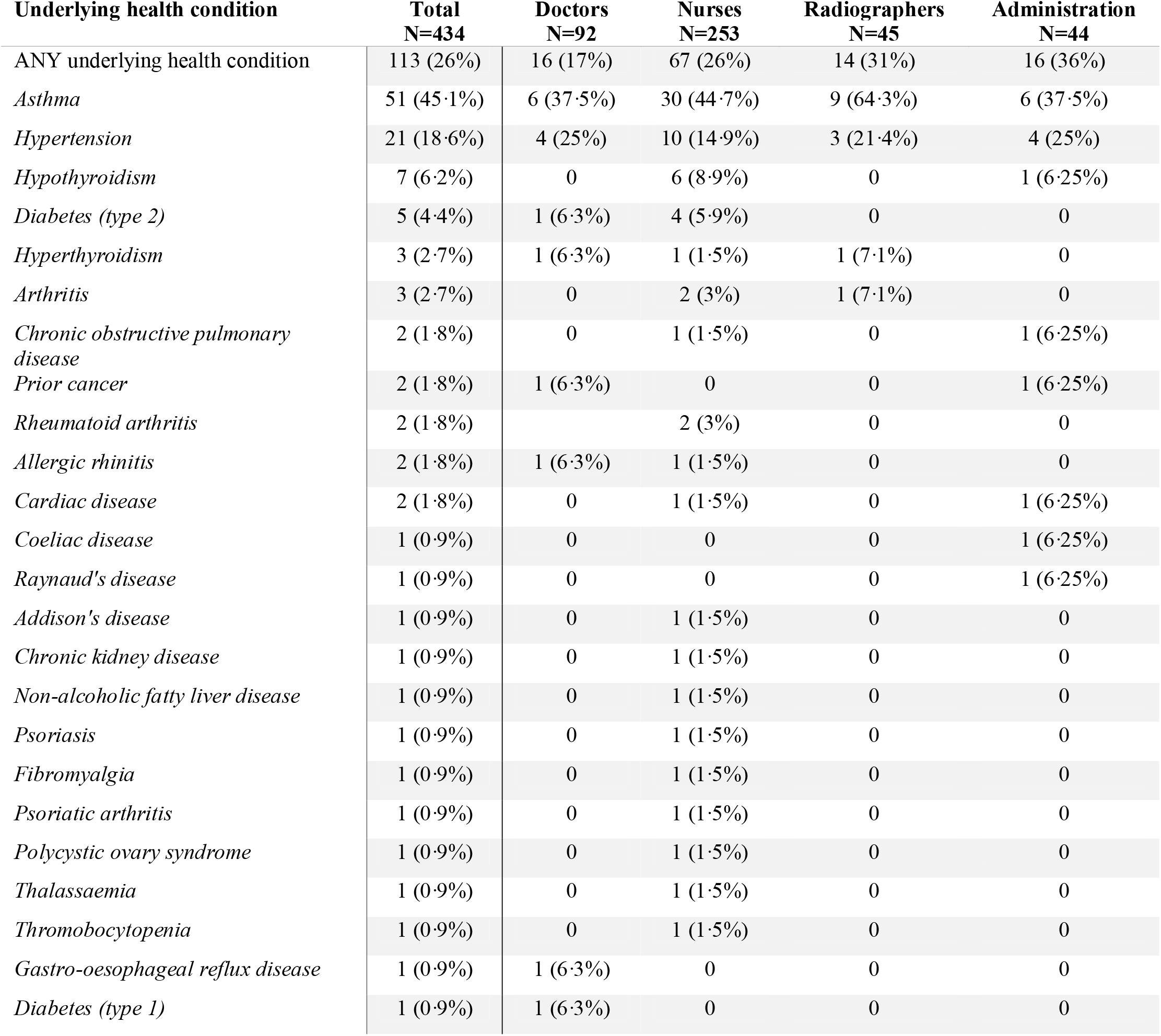
Incidence of reported underlying health conditions in study population. (Some participants had more than one underlying health condition).

**Supplementary Table 2.**
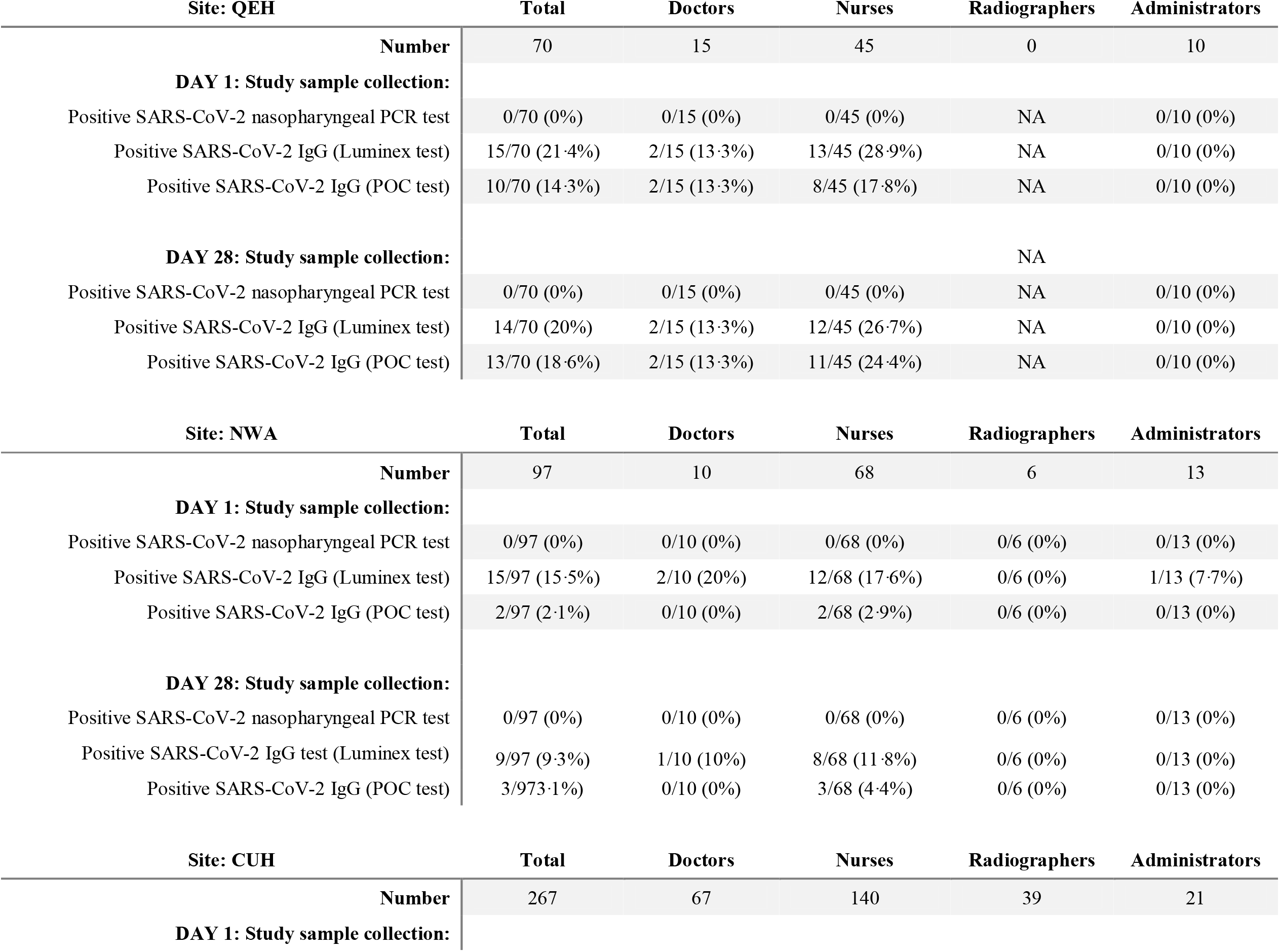
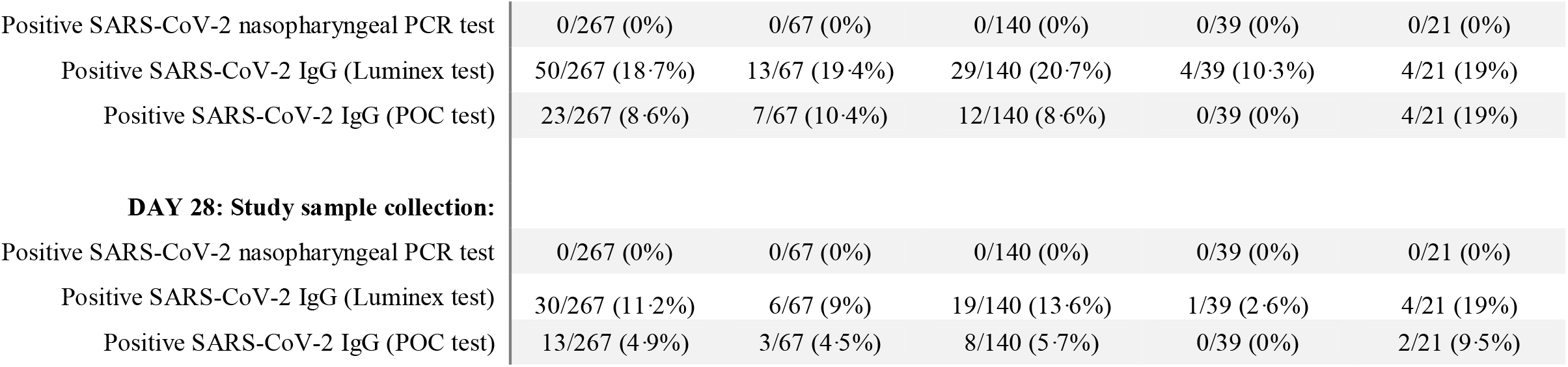
Results by participating hospital site. (NA – not applicable)

| Site: QEH |  | Total | Doctors | Nurses | Radiographers | Administrators |
| --- | --- | --- | --- | --- | --- | --- |
| Number |  | 70 | 15 | 45 | 0 | 10 |
| DAY 1: Study sample collection: |  |  |  |  |  |  |
| Positive SARS-CoV-2 nasopharyngeal PCR test |  | 0/70 (0%) | 0/15 (0%) | 0/45 (0%) | NA | 0/10 (0%) |
| Positive SARS-CoV-2 IgG (Luminex test) |  | 15/70 (21.4%) | 2/15 (13.3%) | 13/45 (28.9%) | NA | 0/10 (0%) |
| Positive SARS-CoV-2 IgG (POC test) |  | 10/70 (14.3%) | 2/15 (13.3%) | 8/45 (17.8%) | NA | 0/10 (0%) |
| DAY 28: Study sample collection: |  |  |  |  | NA |  |
| Positive SARS-CoV-2 nasopharyngeal PCR test |  | 0/70 (0%) | 0/15 (0%) | 0/45 (0%) | NA | 0/10 (0%) |
| Positive SARS-CoV-2 IgG (Luminex test) |  | 14/70 (20%) | 2/15 (13.3%) | 12/45 (26.7%) | NA | 0/10 (0%) |
| Positive SARS-CoV-2 IgG (POC test) |  | 13/70 (18.6%) | 2/15 (13.3%) | 11/45 (24.4%) | NA | 0/10 (0%) |
| Site: NWA |  | Total | Doctors | Nurses | Radiographers | Administrators |
| Number |  | 97 | 10 | 68 | 6 | 13 |
| DAY 1: Study sample collection: |  |  |  |  |  |  |
| Positive SARS-CoV-2 nasopharyngeal PCR test |  | 0/97 (0%) | 0/10 (0%) | 0/68 (0%) | 0/6 (0%) | 0/13 (0%) |
| Positive SARS-CoV-2 IgG (Luminex test) |  | 15/97 (15.5%) | 2/10 (20%) | 12/68 (17.6%) | 0/6 (0%) | 1/13 (7.7%) |
| Positive SARS-CoV-2 IgG (POC test) |  | 2/97 (2.1%) | 0/10 (0%) | 2/68 (2.9%) | 0/6 (0%) | 0/13 (0%) |
| DAY 28: Study sample collection: |  |  |  |  |  |  |
| Positive SARS-CoV-2 nasopharyngeal PCR test |  | 0/97 (0%) | 0/10 (0%) | 0/68 (0%) | 0/6 (0%) | 0/13 (0%) |
| Positive SARS-CoV-2 IgG test (Luminex test) |  | 9/97 (9.3%) | 1/10 (10%) | 8/68 (11.8%) | 0/6 (0%) | 0/13 (0%) |
| Positive SARS-CoV-2 IgG (POC test) |  | 3/97 (3.1%) | 0/10 (0%) | 3/68 (4.4%) | 0/6 (0%) | 0/13 (0%) |
| Site: CUH |  | Total | Doctors | Nurses | Radiographers | Administrators |
| Number |  | 267 | 67 | 140 | 39 | 21 |
| DAY 1: Study sample collection: |  |  |  |  |  |  |

|  |  |  |  |  |  |
| --- | --- | --- | --- | --- | --- |
| Positive SARS-CoV-2 nasopharyngeal PCR test | 0/267 (0%) | 0/67 (0%) | 0/140 (0%) | 0/39 (0%) | 0/21 (0%) |
| Positive SARS-CoV-2 IgG (Luminex test) | 50/267 (18.7%) | 13/67 (19.4%) | 29/140 (20.7%) | 4/39 (10.3%) | 4/21 (19%) |
| Positive SARS-CoV-2 IgG (POC test) | 23/267 (8.6%) | 7/67 (10.4%) | 12/140 (8.6%) | 0/39 (0%) | 4/21 (19%) |
| **DAY 28: Study sample collection:** |  |  |  |  |  |
| Positive SARS-CoV-2 nasopharyngeal PCR test | 0/267 (0%) | 0/67 (0%) | 0/140 (0%) | 0/39 (0%) | 0/21 (0%) |
| Positive SARS-CoV-2 IgG (Luminex test) | 30/267 (11.2%) | 6/67 (9%) | 19/140 (13.6%) | 1/39 (2.6%) | 4/21 (19%) |
| Positive SARS-CoV-2 IgG (POC test) | 13/267 (4.9%) | 3/67 (4.5%) | 8/140 (5.7%) | 0/39 (0%) | 2/21 (9.5%) |

**Supplementary Table 3.** Level of detection comparison between SARS-CoV-2 assays.

|  | POC test |  | Luminex test |  |
| --- | --- | --- | --- | --- |
|  | Positive sample | Negative sample | Positive sample | Negative sample |
| **No dilution** | **positive** | negative | **positive** | negative |
| **1:100** | negative | negative | **positive** | negative |
| **1:250** | negative | negative | negative | negative |
| **1:500** | negative | negative | negative | negative |
| **1:1000** | negative | negative | negative | negative |
| **1:1500** | negative | negative | negative | negative |

**Supplementary Table 4.** Antigenic targets in SARS-CoV-2 IgG antibody-positive participants contrasted by presence or absence of reported symptoms. *Columns listing frequency of changes to original positive cohort: loss of positive antibody status; loss to follow-up; new seroconversions; and change in antibody epitope from IgG targeting SARS-CoV-2 N antigen to S antigen· (N-nucleocapsid; S-surface).

|  | **Antibody**<br>**epitope** | **Day 1**<br>**(positive)** | **Loss of**<br>**positivity\*** | **Lost to**<br>**follow-up\*** | **New sero-**<br>**conversions\*** | **Epitope**<br>**changes\*** | **Day 28**<br>**(positive)** |
| --- | --- | --- | --- | --- | --- | --- | --- |
| **Previously reported symptoms** | N and S antigens | **24** | 0 | 2 | 0 |  | **22** |
|  | N antigen only | **3** | 0 | 1 | 0 | 1x N->S swap | **1** |
|  | S antigen only | **3** | 1 | 0 | 1 |  | **4** |
| **Previously asymptomatic** | N and S antigens | **14** | 3 | 0 | 1 |  | **12** |
|  | N antigen only | **29** | 18 | 2 | 0 | 1x N->S swap | **8** |
|  | S antigen only | **7** | 4 | 0 | 2 |  | **6** |
|  | **Total** | **80** | **-26** | **-5** | **+4** |  | **53** |

